# Outbreak of High-risk XDR CRAB of International Clone 2 (IC2) in Rio Janeiro, Brazil

**DOI:** 10.1101/2023.04.27.23289201

**Authors:** Sérgio M. Morgado, Érica L. Fonseca, Fernanda S. Freitas, Nathalia S. Bighi, Priscila P. C. Oliveira, Priscilla M. Monteiro, Lorena S. Lima, Bianca P. Santos, Maria A. R. Sousa, Adriana O. Assumpção, Luiz A. Mascarenhas, Ana Carolina P. Vicente

## Abstract

The high-risk clones of *Acinetobacter baumannii*, called international clones (ICs), span several ICs, IC1 to IC7 being the most reported. Among them, IC2 represents the main lineage causing outbreaks worldwide. IC2 presents a great diversity of sub-lineages with different sequence types (ST). Despite successful global spread, the occurrence of IC2 is rarely reported in Latin America, including Brazil. Here, through genomic analysis, we investigated 16 IC2/ST2 strains obtained from a clinical setting in Brazil (2022). These strains represented carbapenem-resistant *A. baumannii* (CRAB) with an extensively drug-resistant (XDR) profile, with most showing some resistance to tigecycline. Overall, these strains showed susceptibility only to polymyxins, thus leading to restricted therapeutic choices. Based on *in silico* analysis, the relationship between Brazilian CRAB genomes and IC2/ST2 genomes circulating in the world, particularly in South America, was established. The Brazilian IC2 strains belonged to three discrete and distinct sub-lineages, being more associated with IC2/ST2 genomes from countries in Europe, North America and Asia, suggesting their epidemiological link. These Brazilian strains were characterized by the co-presence of *bla*OXA-23 and *bla*OXA-66, in addition to the genes APH(6), APH(3”), ANT(3”), AAC(6’), *arm*A, and the efflux pumps *ade*ABC and *ade*IJK, associated with the XDR/CRAB phenotype. Furthermore, the three sub-lineages presented three distinct capsules, KL7, KL9 and KL56, the latter not yet associated with IC2 genomes. A large set of virulence genes was also identified in the genomes: *ade*FGH/efflux pump, the siderophores *bar*AB, *bas*ABCDFGHIJ and *bau*BCDEF, *lpx*ABCDLM/capsule, *tss*ABCDEFGIKLM/T6SS, *pga*ABCD/biofilm.

## 1. Introduction

*Acinetobacter baumannii* represents a critically opportunistic pathogen worldwide, with rapid dissemination in clinical settings, being associated with various types of infections [1]. This species is a member of the ESKAPE group (*Enterococcus faecium, Staphylococcus aureus, Klebsiella pneumoniae, Acinetobacter baumannii, Pseudomonas aeruginosa*, and *Enterobacter* spp.), which represents a global threat to human health and poses therapeutic challenges due to the increasing resistance profile [2]. Since 2018, carbapenem-resistant *A. baumannii* (CRAB) is considered by the WHO as the number one priority organism for the research and development of antibiotics [3].

High-risk *A. baumannii* lineages, named International Clones (ICs), usually have multidrug (MDR), extensively drug-resistant (XDR), and carbapenem-resistant (CRAB) characteristics [4]. Most isolates identified worldwide span six ICs (IC1 to IC6), where IC2 represents the most prevalent CRAB, occurring endemically and causing outbreaks. Within IC2, there is a great diversity of sub-lineages with different sequence types (ST), with ST2 (Pasteur scheme) being the most prevalent [5]. Indeed, *A. baumannii* presents high levels of recombination events, resulting in distinct lineages, even within the same ST [1]. The presence of divergent IC2 strains and their increasing prevalence in clinical settings support the continued adaptation of this lineage to the hospital environment [6]. However, despite the successful global spread of IC2, its occurrence has barely been reported in Latin America [5,7,8]. In Brazil, *A. baumannii* of IC1, IC4, and IC5 and, to a lesser extent, IC7, have been the most prevalent clones isolated and causing outbreaks throughout the country. This scenario may be evolving since an outbreak of CRAB IC2, characterized by ArmA- and OXA-23-producing strains, was reported in the southeast region (city of São Paulo) [9].

The virulence of *A. baumannii* is multifactorial, determined by the combination of multiple factors such as efflux pumps (*ade*RS), quorum sensing (*aba*), siderophores (*bar, bas, bau*), biofilm (*bap*), secretion systems (*pil*, TXSS), and one of the most important factors, the capsule [10].

Here, we investigated the genomic relatedness of 16 new Brazilian IC2/ST2 genomes from an outbreak in a tertiary hospital in the southeast region (city of Rio de Janeiro). It has been revealed that at least three XDR CRAB IC2/ST2 sub-lineages are causing outbreaks in Brazil since the period of the COVID-19 pandemic. They were characterized by an enormous apparatus of genes associated with antibiotic resistance and virulence, distributed both in mobile elements and in the chromosome.

## 2. Methodology

### 2.1. Isolates and Antimicrobial Resistance Analysis

Sixteen strains of *A. baumannii* were recovered from nosocomial infections that occurred in a tertiary hospital located in Rio de Janeiro (Brazil). Antimicrobial susceptibility testing by the disk-diffusion method was performed for all antibiotics considered for *Acinetobacter* spp. resistance classification [11] and interpreted according to the Clinical and Laboratory Standards Institute (CLSI) [12]. The Minimum Inhibitory Concentration (MIC) of polymyxin B and colistin was determined by the broth microdilution method and interpreted according to the European Committee for Antimicrobial Susceptibility Testing (EUCAST) guidelines (MIC breakpoint for resistance > 2 mg/L [13].

### 2.2. Public Data Set

Complete and draft genomes of *Acinetobacter baumannii* available from Genbank (n = 17,444) were obtained from the National Center for Biotechnology Information (NCBI) on December 2022 (https://ftp.ncbi.nlm.nih.gov/genomes/genbank/bacteria/Acinetobacter_baumannii/).

### 2.3. Genome Sequencing and Assembly

In this study, we generated 16 genomes of nosocomial *Acinetobacter baumannii*. The genomic DNA of these 16 isolates was obtained with NucleoSpin Microbial DNA kit (Macherey-Nagel), and used for library construction with Nextera paired-end library kit. The sequencing was performed on Illumina Hiseq 2500, generating 150 bp-length reads. The raw reads were filtered using NGS QC Toolkit v.2.3.3 (Phred quality score ≥ 20) [14]. The genomes were *de novo* assembled with SPAdes assembler v3.15.2 [15] and polished with Pilon v1.24 [16]. The sequences are available at NCBI bioproject PRJNA938102.

### 2.4. Characterization and Phylogeny of *Acinetobacter baumannii* Genomes

The genomes were annotated with Prokka v1.14.6 [17], and their sequence type was determined with mlst v2.22.0 (https://github.com/tseemann/mlst) using Pasteur scheme. Virulence and antibiotic resistance genes were assayed with ABRicate v1.0.1 (https://github.com/tseemann/abricate) using VFDB and CARD databases, respectively. The capsular polysaccharide (KL) was obtained by Kaptive v2.0 [18]. Figures of gene clusters were drawn using Clinker v0.0.24 [19].

As thousands of IC2/ST2 genomes are available, the phylogeny was performed with genomes more closely related to the genomes sequenced in this study. Thus, the entire genome dataset was indexed with mash software [20], and a subset of those most closely related was selected for further analysis, in addition to genomes from Latin America. The core genome of this subset was estimated using Roary v3.13.0 [21], and sites with single nucleotide polymorphisms (SNPs) were extracted from the concatenated core genes using snp-sites v2.5.1 [22], which were submitted to phylogenetic analysis by IQTree v1.6.12 [23], using the best-fit substitution model (SYM+ASC+R4), to obtain a maximum likelihood tree with 1000 ultrafast bootstrap replicates [24]. The tree was drawn using iTOL web platform (https://itol.embl.de) [25].

## 3. Results

*A. baumannii* IC2/ST2 is considered the most widespread IC globally, however, until now, this IC has only been occasionally reported in South America. Here, we investigated the genomic relationship of 16 new Brazilian IC2/ST2 genomes obtained from a clinical setting in Brazil and, in this way, these were compared with genomes circulating in Latin America, particularly South America, and genomes from the rest of the world.

Of the 17,444 genomes of *A. baumannii* deposited in Genbank, 611 (∼3%) were from Latin America, covering 15 countries (Table 1). These 611 genomes from Latin America belong to the following STs (Pasteur scheme): ST2 (n=105), ST25 (n=64), ST1 (n=38), ST156 (n=35), ST79 (n=26), ST15 (n=21), ST991 (n=18). However, regarding IC2/ST2, most Latin American countries are underrepresented, as 91/105 are from a single country (Mexico), while Brazil presented only one IC2/ST2 genome. The other genomes of *A. baumannii* from Brazil belonged to IC5/ST79 (n=78), IC1/ST1 (n=42), ST15 (n=32) and IC7/ST25 (n=18).

### 3.1. *A. baumannii* IC2/ST2 resistome

All 16 clinical IC2/ST2 strains from an outbreak in southeastern Brazil (Rio de Janeiro, 2022) showed the XDR phenotype (Table 2), being susceptible only to polymyxins. In fact, the analysis of the *in silico* resistome revealed the presence of genes associated with resistance to fluoroquinolones (AAC(6’)-Ib7), carbapenems (ADC-30, OXA-23, OXA-66), aminoglycosides (ANT(3”)-IIa, APH(3’’)-Ib, APH(6)-Id, *arm*A), phenicol (*cat*B8), macrolides (*mph*E, *msr*E), sulfonamides (*sul*1), and tetracyclines (*tet*(B)) (Table 2; Table S1). In addition, we were able to map, in all Brazilian genomes, a resistance island similar to AbaR4 (∼23 kb in size), which harbored *tet*(B), OXA-23, APH(3’’)-Ib, and APH(6)-Id, and it was flanked by sequences from the *com*M gene. In fact, most AbaR islands use this insertion site [26]. This island showed ∼100% coverage to the AB210 genome island (HQ700358.2) (England/2005). Furthermore, in addition to OXA-23 in the resistance island, all Brazilian genomes presented a blaOXA-66 in the chromosome.

In addition to the acquired resistome, all 16 Brazilian genomes showed amino acid substitutions in GyrA (S81L) and ParC (S84L, V104I, and D105E), which are associated with resistance to fluoroquinolones. In addition, they encoded several multidrug efflux complexes such as AbaF (associated with fosfomycin resistance and biofilm formation), AdeABC (associated with glycylcycline, aminoglycosides, β-lactams, chloramphenicol, erythromycin, fluoroquinolones, tetracycline, trimethoprim resistance), AdeIJK (associated with carbapenem, chloramphenicol, erythromycin, fluoroquinolone, lincosamides, rifampicin, tetracycline, trimethoprim resistance), AdeFGH (associated with fluoroquinolone and tetracycline resistance). Considering these intrinsic drug efflux pumps, all 16 genomes presented gene sequences with 100% identity among themselves, including the regulatory genes *ade*R and *ade*S. The exceptions were *aba*F of Ab115, which had a mutation (C410A) resulting in a stop codon; and *ade*G from Ab114 with a non-synonymous mutation (G896T -> G299V). Furthermore, the AdeR and AdeS sequences were compared with reference sequences from *A. baumannii* ATCC 17978 (CP053098.1) to identify possible amino acid substitutions. We observed, in all 16 genomes, the substitutions of V120I and A136V in AdeR; and I62M, L172P, G186V, N268H, Y303F, and V348I in AdeS.

Regarding the other IC2 genomes analyzed (n=103), all but three were also characterized by chromosomal OXA-66, most in combination with OXA-23. The other genomes showed combinations of OXA-66/OXA-72, OXA-66/OXA-237, OXA-66/OXA-239, OXA-66/OXA-398 and OXA-66/OXA-657 (Table S1). *In silico* analysis of the resistome of these other genomes also revealed a high prevalence of some genes associated with the mobilome: AAC(6’)-Ib7, ADC-30, ANT(3’’)-IIa, APH(3’’)-Ib, APH(6)-Id, *arm*A, and *cat*B8 (Figure 1).

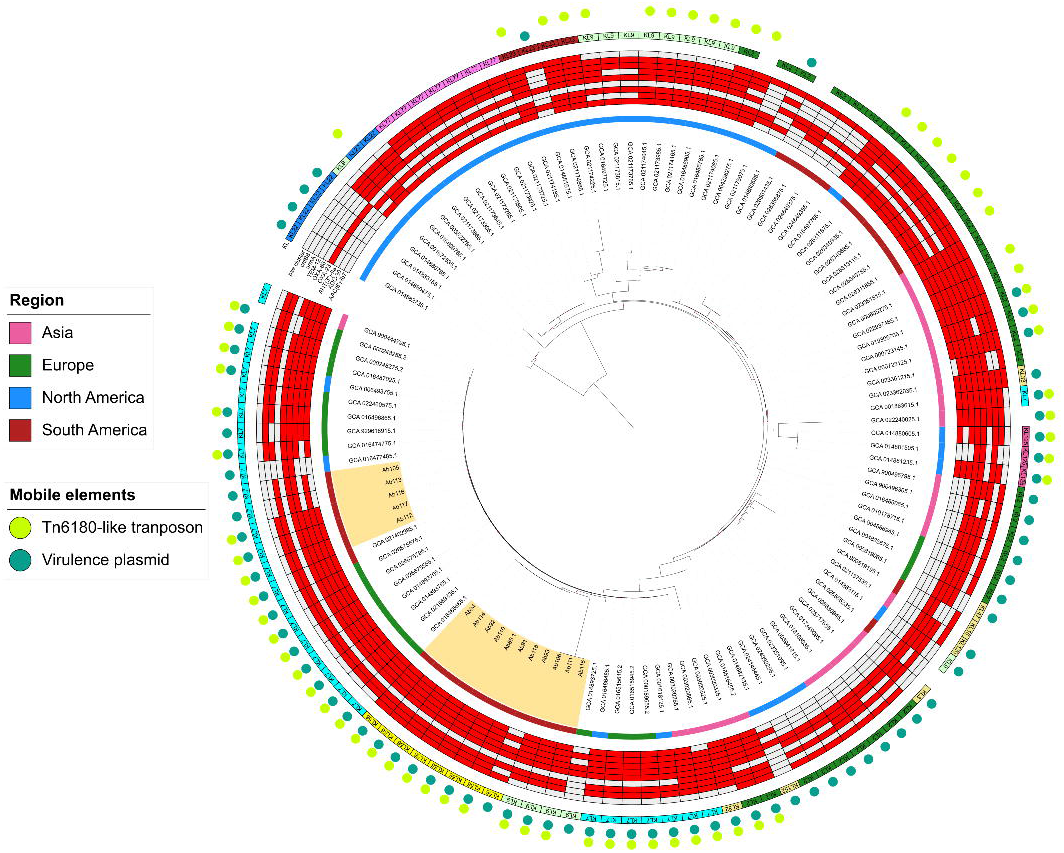

### 3.2. *A. baumannii* IC2/ST2 virulome

Few genomes of other KL (KL33, KL213, KL16, KL120, KL56) also harbored *pse*BCFI genes sharing high identity (>90%) with equivalent genes in KL2. However, KL120 and KL56 presented the *pse*G (*psa*D)/*pse*H (*psa*E) region replaced by *psa*G/*psa*H genes, which have low identity to *pse*G (*psa*D) and *pse*H (*psaE*) (Figure 2).

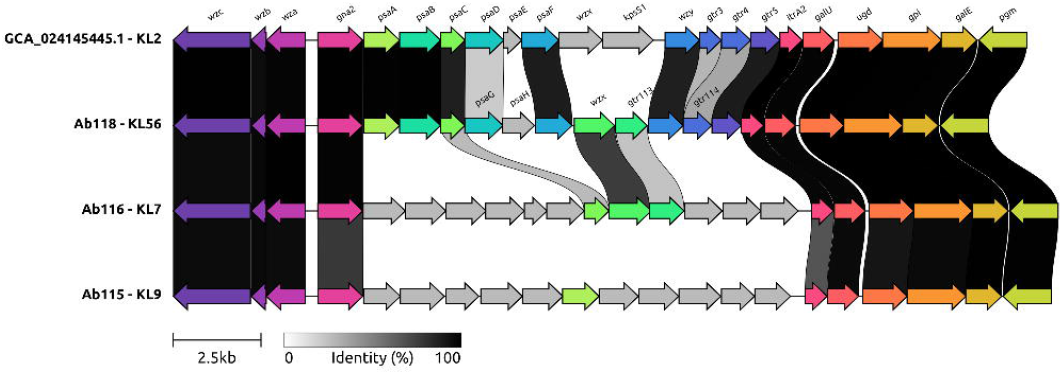

The virulome was also vast and appears to be homogeneous in this set of 119 IC2/ST2 genomes (efflux pump, *ade*FGH; siderophores, *bar*AB, *bas*ABCDFGHIJ, and *bau*BCDEF; capsule-associated genes, *lpx*ABCDLM; T6SS, *tss*ABCDEFGIKLM; biofilm, *pga*ABCD; etc), except by the absence/presence of few genes, in some genomes, related to gene clusters of the Type VI secretion system and capsule (Table S2). Most genomes carried KL2, but KL7, KL9, KL22, KL23, KL77, and KL125 were represented in several genomes (Figure 1). The 16 Brazilian genomes presented three different capsular polysaccharides (KL): KL7 (n=5), KL9 (n=3), and KL56 (n=8) (Figure 2). Interestingly, KL56 had not yet been associated with any IC2/ST2 (even considering the entire IC2/ST2 dataset). Among the virulence genes considered, the *pse* gene cluster, *pse*B (*psa*A), *pse*C (*psa*B), *pse*F (*psa*C), *pse*G (*psa*D), *pse*H (*psa*E), *pse*I (*psa*F), which encodes the pseudaminic acid (associated with the capsule apparatus), was identified only in part of the analyzed genomes, being present mainly in the KL2, KL77 and KL23 genomes (Table S2). Few other KL genomes (KL33, KL213, KL16, KL120, KL56) also harbored *pse*BCFI genes sharing high identity (>90%) with equivalent genes in KL2. However, KL120 and KL56 presented the *pse*G (*psa*D)/*pse*H (*psa*E) region replaced by the *psa*G/*psa*H genes, which have a low identity with *pse*G (*psa*D) and *pse*H (*psa*E) (Figure 2).

## 3.3. *A. baumannii* IC2/ST2 mobilome

The mobilome of all 16 Brazilian genomes was analyzed to identify the context and origin of some genes. Two contigs carrying *rep* genes were identified in most 13/16 genomes (Figure 1). One contig carried antibiotic resistance genes (ARG) and the other, virulence genes. The ARG-bearing contig was

∼17 kb in size and harbored the genes repM_Acin (RepB family), *arm*A, *mph*E, *msr*E, AAC(6’)-Ib7, ANT(3’’)-IIa, *cat*B8, *sul*1, *qac*Edelta1, the last five being part of a class I integron. In addition, several insertion sequences (ISs) were observed, such as IS66-like (ISAba24), IS4-like, and IS5. This contig showed 100% identity and ∼89% coverage with plasmid p3BJAB0868 (CP003908.1; China/2007); 100% identity and ∼96% coverage (missing an IS26) with the Tn1548-like-1 transposon (KT317079.1; Sweden/2012); and 100% identity and ∼93% coverage (missing two IS26s) with the Tn6180 transposon, which is embedded in the AbGRI3 resistance island (KX011025.2; Singapore/2007) (Figure 3). The lack of information from the ISs could be because the ISs are associated with repetitive sequences, therefore, they could not have been assembled in this contig ARG. In fact, through mapping, these transposons were obtained completely. Therefore, this element seems to be the main contributor to the acquired resistome of the Brazilian XDR CRABs. The contig carrying virulence genes resembles a small plasmid (∼8.8 kb), characterized by a repM_Acin (RepB family), and with ∼100% identity and coverage with plasmid p1BJAB0868 (CP003850.1; China/2007), being present in all 16 genomes. It encoded a toxin (BrnT)/antitoxin (BrnA) system, septicolysin, *sel*1 gene, and TonB-dependent receptor genes. Furthermore, mobile elements similar to those two described (considering sequence coverage > 70%) were also observed in several other IC2/ST2 genomes from the analyzed dataset (Figure 1). Interestingly, another plasmid with a size of ∼66 kb, characterized by a *rep*A, was identified in only one genome, Ab115. It presented a set of genes resembling the type IV secretion system (T4SS), in addition to a relaxase gene, thus conferring a conjugative character to this plasmid. It showed ∼100% identity and coverage to the plasmid pCRA914-67 (MK243454.1; Canada). In addition to the conjugative apparatus, this plasmid also harbored a toxin (RelE-like)/antitoxin (Xre-like) system, Zeta toxin, *par*A/*par*B partitioning genes, and *tel*A gene (tellurite and oxyanions resistance).

### 3.4. *A. baumannii* IC2/ST2 phylogeny

Phylogenetic analysis showed that the 16 Brazilian genomes from the city of Rio de Janeiro were clustered into three distinct and discrete groups, in which two groups contained only Brazilian genomes (Figure 1). The third group contained 5/16 of the genomes sequenced here clustered with the previously unique Brazilian IC2/ST2 genome (GCA_021402995.1; from the city of São Paulo) and next to genomes from North America, Europe, and Asia. Each of these three groups was characterized by a capsular polysaccharide locus, with KL7 and KL9 also being present in other genomes, and KL56 occurring only in this set of IC2/ST2 genomes from Rio de Janeiro. Of note, no IC2/ST2 genomes from Latin America belonged to groups containing Brazilian genomes, therefore current IC2/ST2 circulating in Brazilian clinical settings is more likely to be associated with IC2/ST2 lineages occurring in Europe, North America and/or Asia.

## 4. Discussion

CRAB IC2 is the most prevalent *A. baumannii* causing outbreaks worldwide [8], however, interestingly, reports of IC2/ST2 are scarce in South America [7]. In fact, in a recent study (2022) on the origin, phylogeny, and transmission of IC2 on a global scale, IC2 from South America was not analyzed [1]. Furthermore, the study concluded that IC2 (ST208/Oxford scheme) may have originated in North America and subsequently evolved into two clades [1]. Interestingly, in the same year (2022), the first CRAB IC2 outbreak was reported in Brazil (São Paulo city), and an IC2/ST2 genome from Brazil became available [9]. In the same year (2022), we made available dozens of new CRAB IC2/ST2 genomes from an outbreak in the city of Rio de Janeiro, providing important epidemiological data on this supposedly rare IC in South America.

The IC2/ST2 genome from the city of São Paulo (GCA_021402995.1) showed a vast resistome, including genes associated with resistance to aminoglycosides, carbapenems, and fluoroquinolones, among others. Its resistome was in agreement with the resistance phenotype of the strain [9]. Likewise, the same was observed within the set of CRAB IC2/ST2 from Rio de Janeiro, in agreement with the resistance phenotype (XDR). In general, the strains in the present study were only susceptible to polymyxins, like other IC2/ST2 strains from South America [7,9]. In addition to presenting similar resistance profiles, the genome from the São Paulo outbreak belongs to the same cluster as the Rio de Janeiro genomes, showing that this XDR CRAB IC2/ST2 sub-lineage is currently widespread in at least two states in the southeast region of Brazil. Interestingly, in this same cluster, characterized by the genomes with the KL7 capsule, there were also genomes from Portugal and the United Kingdom, recovered in 2012 and 2007, respectively, which shared the same set of resistance and virulence genes. Therefore, this XDR CRAB IC2/ST2 sub-lineage appears to have an enormous potential for dissemination and persistence, since it has been circulating in the world for over a decade. The other Brazilian genomes formed two discrete clusters apart from the last cluster, as well as other clusters characterized by global IC2 genomes. Overall, this suggests that there are at least three XDR CRAB IC2/ST2 sub-lineages occurring in Brazil simultaneously. Interestingly, the Brazilian genomes did not cluster with other IC2/ST2 genomes from South America, suggesting that these Brazilian CRAB IC2/ST2 strains originate from countries in Europe, North America, and/or Asia where CRAB IC2 is prevalent [1]. Although we did not observe relevant differences in the genetic composition of these three Brazilian clusters, except for the capsule, they could be evolving into specific lineages, as seems to have occurred for other South American ICs [27].

The remarkable characteristic of antibiotic resistance of *A. baumannii* is mainly attributed to resistance islands associated with transposons (AbaR) and their variations (AbaR-like regions), usually inserted in the *com*M gene [26]. Indeed, the genomes sequenced here carried most of the antibiotic resistance genes in elements such as Tn6180-like and AbaR4-like. The Tn6180-like element also showed a *rep* gene and identity with plasmid p3BJAB0868, which would characterize it as a plasmid. However, this *rep* gene had 100% identity/coverage with a *rep* that would be truncated, thus being non-functional [28]. Thus, it is likely that this element behaves like a transposon. This issue has already been raised in other studies, as this transposon and related ones could assume a circular shape, like a plasmid [29,30].

The ARG and virulence mobile elements identified here appear to be scattered among IC2 *A. baumannii*, as observed in several other genomes in the current dataset and also reported in IC2 genomes from India [31] and China [32]. In addition, virulence and conjugative plasmids have also been reported in other ICs, such as IC5 and IC7, in Bolivia [33], a country that borders Brazil. Considering antibiotic resistance by efflux pumps, we observed that the *ade*ABC regulatory genes, *ade*RS, encoded proteins with some amino acid substitutions. Among them, A136V in AdeR and G186V in AdeS have been associated with overexpression of the AdeABC efflux system and lower susceptibility to ciprofloxacin, gentamicin, and tigecycline resistance [34-36]. In fact, most of the strains in this study showed the resistance phenotype to ciprofloxacin and gentamicin (Table 2). Thus, the Brazilian genomes, in addition to having a mobile resistome (susceptible to frequent gains/losses), also have intrinsic and more stable resistance mechanisms.

Among the virulence genes identified, the *pse* genes, which encode pseudaminic acid, were present only in some genomes of specific KL (KL2, KL77, KL23, KL33, KL213, KL16, KL120, KL56). Pseudaminic acids are non-mammalian acid sugars present in glycoconjugates on the surface of pathogenic bacteria [37]. It is a rare sugar in the capsule of *A. baumannii* and its presence has been correlated with enhanced virulence [38]. This acid is required for flagellar assembly and motility in *Campylobacter jejuni* and *Helicobacter pylori*, and is also involved with autoagglutination, adhesion, invasion, and immune evasion [39]. Since *A. baumannii* does not have flagella, exhibiting twitching motility [40], the role of pseudaminic acid should be associated with functions other than motility. As the Brazilian genomes carrying the Pse cluster (KL56) had the *pse*G (*psa*D)/*pse*H (*psa*E) region replaced by the *psa*G/*psa*H genes, the processing of substrates in the pseudaminic acid pathway must be different in these organisms [41].

Therefore, this study revealed that widespread high-risk XDR CRAB IC2/ST2 is currently causing outbreaks in clinical settings in the southeast region. As these difficult-to-control strains are only susceptible to polymyxin, therapeutic options are limited and there is a risk of developing pan-resistance. Furthermore, this is due to at least three sub-lineages characterized by an enormous apparatus of virulence and resistance to antibiotics, both intrinsic and mobile.

## Data Availability

All data produced in the present work are contained in the manuscript

## Ethical approval

Not required.

---

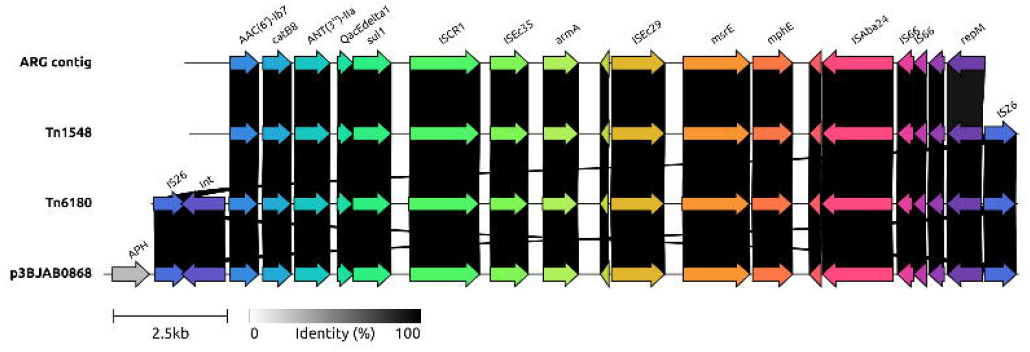

