## Supplementary material for "Outbreak of High-risk XDR CRAB of International Clone 2 (IC2) in Rio Janeiro, Brazil": Figure

**Figure legends**

**Figure 1. Maximum likelihood tree based on the core genome of IC2/ST2 *A. baumannii*.** The innermost orbit of the colored blocks represents the geographic region where the genome was obtained. The red (identified) and gray (unidentified) blocks represent the identification or not of such a gene in that genome. The outermost orbit of the colored blocks represents the capsule locus (KL) of this genome. The outer circles mark the genomes with the virulence plasmid and the ARG transposon. The genomes sequenced in this study are shaded in yellow. The red circles on the branches represent bootstrap >70%.

**Figure 2. *A. baumannii* capsule gene cluster.** Comparison of the capsule region of the Brazilian genomes of IC2/ST2 A. baumannii and reference KL2. Arrows represent predicted genes, which are colored based on their similarity. Gene identity between genomes is shown by grayscale links.

**Figure 3. *A. baumannii* transposon gene cluster.** Comparison of the transposon region carrying antibiotic resistance genes from *A. baumannii* IC2/ST2 genomes. Arrows represent predicted genes, which are colored based on their similarity. Gene identity between regions is shown by grayscale links.
