## Supplementary material for "Outbreak of High-risk XDR CRAB of International Clone 2 (IC2) in Rio Janeiro, Brazil": Table

Table 1. Number of *A. baumannii* genomes in Latin America

| Country | # Number of genomes | # Number of ST2 genomes |
| --- | --- | --- |
| Argentina | 39 | 4 |
| Bolivia | 55 | 0 |
| Brazil | 229 | 1 |
| Chile | 49 | 0 |
| Colombia | 16 | 0 |
| Dominican Republic | 9 | 0 |
| Ecuador | 8 | 6 |
| El Salvador | 2 | 0 |
| Honduras | 20 | 0 |
| Mexico | 134 | 91 |
| Nicaragua | 2 | 0 |
| Paraguay | 37 | 1 |
| Peru | 4 | 0 |
| Puerto Rico | 2 | 1 |
| Venezuela | 5 | 1 |

Table 2. Antibiotic susceptibility of the Brazilian A. baumannii

| Antibiotic classes | Antibiotics | Strains | | | | | | | | | | | | | | | | Genes associated | |
| --- | --- | --- | --- | --- | --- | --- | --- | --- | --- | --- | --- | --- | --- | --- | --- | --- | --- | --- | --- |
|  |  | Ab85.1 | Ab91 | Ab92 | Ab93 | Ab94 | Ab105 | Ab106 | Ab110 | Ab111 | Ab112 | Ab113 | Ab114 | Ab115 | Ab116 | Ab117 | Ab118 | Chromosomic | Mobile |
| Aminoglycosides | AMK | R | R | S | S | R | R | R | R | R | R | R | R | R | R | R | R | *ade*ABC | APH(6); APH(3''); ANT(3''); AAC(6'); *arm*A |
|  | GEN | R | R | S | S | R | R | R | R | R | R | R | R | R | R | R | R |  |  |
|  | NET | R | R | R | S | R | R | R | R | R | R | R | R | R | R | R | R |  |  |
|  | TOB | R | R | S | S | R | R | R | R | R | R | R | R | R | R | R | R |  |  |
| Carbapenems | IPM | R | R | R | R | R | R | R | R | R | R | R | R | R | R | R | R | *ade*ABC; *ade*IJK; OXA-66 | OXA-23 |
|  | MEM | R | R | R | R | R | R | R | R | R | R | R | R | R | R | R | R |  |  |
|  | ETP | R | R | R | R | R | R | R | R | R | R | R | R | R | R | R | R |  |  |
|  | DOR | R | R | R | R | R | R | R | R | R | R | R | R | R | R | R | R |  |  |
| Cephalosporins | CAZ | R | R | R | R | R | R | R | R | R | R | R | R | R | R | R | R | *ade*IJK; OXA-66 | ADC-30; OXA-23 |
|  | CRO | R | R | R | R | R | R | R | R | R | R | R | R | R | R | R | R |  |  |
|  | CTX | R | R | R | R | R | R | R | R | R | R | R | R | R | R | R | R |  |  |
|  | FEP | R | R | R | R | R | R | R | R | R | R | R | R | R | R | R | R |  |  |
| β-lactams | SAM | R | R | R | R | I | R | R | R | R | R | R | R | R | R | R | R | *ade*IJK; *ade*ABC; OXA-66 | ADC-30; OXA-23 |
|  | TZP | R | R | R | R | R | R | R | R | R | R | R | R | R | R | R | R |  |  |
|  | TIM | R | R | R | R | R | R | R | R | R | R | R | R | R | R | R | R |  |  |
| Phenicols | CHL | R | R | R | R | R | R | R | R | R | R | R | R | R | R | R | R | *ade*ABC; *ade*FGH; *ade*IJK | *cat*B8 |
| Quinolones | CIP | R | R | R | R | R | R | R | R | R | R | R | R | R | R | R | R | *ade*FGH; *ade*IJK; *gyr*A/*par*C ; *ade*I; a*ba*Q; *ade*ABC | AAC(6') |
|  | LEV | R | R | R | R | R | R | R | R | I | R | R | R | I | R | R | R |  |  |
| Macrolides | ERY | R | R | R | R | R | R | R | R | R | R | R | R | R | R | R | R | *ade*IJK; *ade*ABC | *mph*E; *msr*E |
| Sulfonamides/Antifolate | SXT | R | R | R | I | R | R | R | R | R | R | R | R | R | R | R | R | *ade*ABC; *ade*IJK | *sul*1 |
| Tetracyclines | MIN | R | R | R | I | R | R | I | R | S | I | I | S | S | R | R | I | *ade*FGH; *ade*IJK; *ade*ABC | *tet*(B); *tet*R |
|  | TET | S | R | R | R | R | R | R | R | R | R | R | R | R | R | R | R |  |  |
|  | DOX | S | R | R | R | R | R | R | R | R | R | R | R | I | R | R | R |  |  |
|  | TGC | R | R | R | S | R | S | I | R | I | S | S | R | I | S | S | R |  |  |

AMK, amikacin; GEN, gentamicin; NET, netilmicin; TOB, tobramycin; IPM, imipenem; MEM, meropenem; ETP, ertapenem; DOR, doripenem; CAZ, ceftazidime; CRO, ceftriaxone; CTX, cefotaxime; FEP, cefepime; SAM, ampicillin/sulbactam; TZP, piperacillin/tazobactam; TIM, ticarcillin/clavulanic acid; CHL, chloramphenicol; CIP, ciprofloxacin; LEV, levofloxacin; ERY, erythromycin; SXT, trimethoprim/sulfamethoxazole; MIN, minocycline; TET, tetracycline; DOX, doxycycline; TGC, tigecycline. Resistance profile: S, susceptible; R, resistant; I, intermediate.
